# Patient-reported Vision Quality-of-life in Parkinsonian Syndromes and Ataxias and Association with Clinical Oculomotor Findings

**DOI:** 10.64898/2026.02.03.26345521

**Authors:** Faye X. Yang, Rohin Manohar, Anna C. Luddy, Albert Y. Hung, Anne-Marie A. Wills, Christopher D. Stephen, Jeremy D. Schmahmann, Anoopum S. Gupta

## Abstract

**Background:** Oculomotor dysfunction is common in parkinsonian syndromes and ataxias, but its impact on patient-reported vision-related quality of life (VQoL) remains insufficiently understood.

**Objectives:** To characterize VQoL across parkinsonian syndromes and ataxias and assess the functional significance of specific oculomotor abnormalities in spinocerebellar ataxias.

**Methods:** Participants were recruited at Massachusetts General Hospital (*n*=231): 104 with Parkinson’s disease (PD), 10 with progressive supranuclear palsy (PSP), 56 with genetically defined ataxias (SCA2, SCA3, SCA6, SCA27B, CANVAS), and 61 healthy controls. VQoL was assessed using a 13-item subset of the Visual Activities Questionnaire targeting depth perception, visual acuity/spatial vision, and visual processing speed. Clinical severity was assessed with the Brief Ataxia Rating Scale and Modified International Cooperative Ataxia Rating Scale, and subjective symptoms with PROM-Ataxia. Group comparisons, correlations, and regression analyses were performed.

**Results:** All disease groups reported significantly worse VQoL than controls, with the largest deficits in visual processing speed. PSP showed the greatest impairment across all domains, while PD was less affected. Individuals with SCA3 and SCA6 had significantly lower VQoL across all subcategories. In ataxias, VQoL correlated moderately with PROM-Ataxia and weakly with clinical oculomotor scores. Gaze-evoked nystagmus was the only oculomotor sign independently associated with reduced VQoL.

**Conclusions:** Parkinsonian syndromes and ataxias are associated with substantial VQoL impairment, particularly in visual processing speed. Gaze-evoked nystagmus is a key predictor of reduced VQoL in ataxias, highlighting the functional relevance of fixation instability. Patient-reported outcomes and oculomotor assessments are essential for capturing visual disability in clinical care and trials.

## Introduction

Declining visual function profoundly affects quality of life by compromising mobility, independence, and social engagement (1–3). Adults with vision loss experience greater difficulty with activities of daily living and increased healthcare utilization compared to peers with intact vision (4–7). While vision loss is commonly attributed to ocular disease, vision-related dysfunction also arises from neurodegenerative conditions (8–10). Parkinsonian syndromes and ataxias are characterized by abnormalities in saccades, pursuit, and fixation, which may interfere with everyday visual tasks (11–14), yet their impact on patient-reported vision-related quality of life (VQoL) remains underexplored.

Individuals with Parkinson’s disease (PD) frequently experience visual disturbances, with over three-quarters reporting at least one complaint (15). Common complaints include blurred vision, diplopia, light sensitivity, and impaired spatial judgment (16–19). Progressive supranuclear palsy (PSP), an atypical parkinsonian syndrome, has characteristic vertical gaze palsy, slowed saccades, and eyelid apraxia (20,21). Visual symptoms such as photophobia, blurred vision, and diplopia often emerge early in PSP and substantially interfere with reading, driving, and ambulation (22–27).

Oculomotor symptoms are also highly prevalent in spinocerebellar ataxias (SCAs), though their manifestations vary by subtype. SCA2 is associated with slowed saccades (28,29), whereas SCA27B is characterized by downbeat nystagmus that causes oscillopsia and impair visual stability (30,31). Individuals with SCAs exhibit impairments in contrast sensitivity, stereo-acuity, and ocular alignment, leading to difficulties with both near and distance vision (32). Visual symptoms may precede other neurological deficits by several years and are associated with reduced VQoL (33,34).

While prior studies have characterized oculomotor deficits in these disorders using clinical scales and quantitative eye tracking (35–39), few have examined how specific abnormalities relate to everyday visual function. We therefore aimed to (1) characterize VQoL deficits across parkinsonian syndromes and ataxias relative to age-matched controls, and (2) identify which clinical oculomotor abnormalities are most strongly associated with reduced VQoL in individuals with ataxia.

## Methods

### Participants

The study protocol was approved by the Massachusetts General Hospital Institutional Review Board (2021P000257). Participants were recruited from Neurobooth, a multimodal digital phenotyping study capturing oculomotor, cognitive, speech, and motor behavior (40). Participants completed quality-of-life questionnaires alongside standardized tasks. Data collection for Neurobooth began in April 2022, and the Visual Activities Questionnaire (VAQ) was added in June 2023. VAQ data collected between June 2023 and February 2025 were analyzed.

For the first analysis, VQoL was compared between disease groups and controls. Controls were screened for neurological and significant ophthalmologic conditions. Participants with diabetic retinopathy, glaucoma, and blindness were excluded. The cohort included 231 unique individuals: 104 with Parkinson’s disease (PD), 10 with progressive supranuclear palsy (PSP), 56 with genetically-defined ataxias, and 61 healthy controls. The ataxia cohort included 6 with SCA2, 24 with SCA2, 12 with SCA6, 7 with SCA27B, and 7 with cerebellar ataxia with neuropathy and vestibular areflexia syndrome (CANVAS). Table 1 shows participant demographics in this analysis. Clinical severity was assessed using the Movement Disorder Society-Unified Parkinson’s Disease Rating Scale Part III (MDS-UPDRS) (41) in PD individuals, PSP Staging System (42) in PSP individuals, and Brief Ataxia Rating Scale (BARS) (43) and Scale for the Assessment and Rating of Ataxia (SARA) (44) in ataxia individuals. Scores were rated in person by neurologists during same-day visits.

**Table 1.**
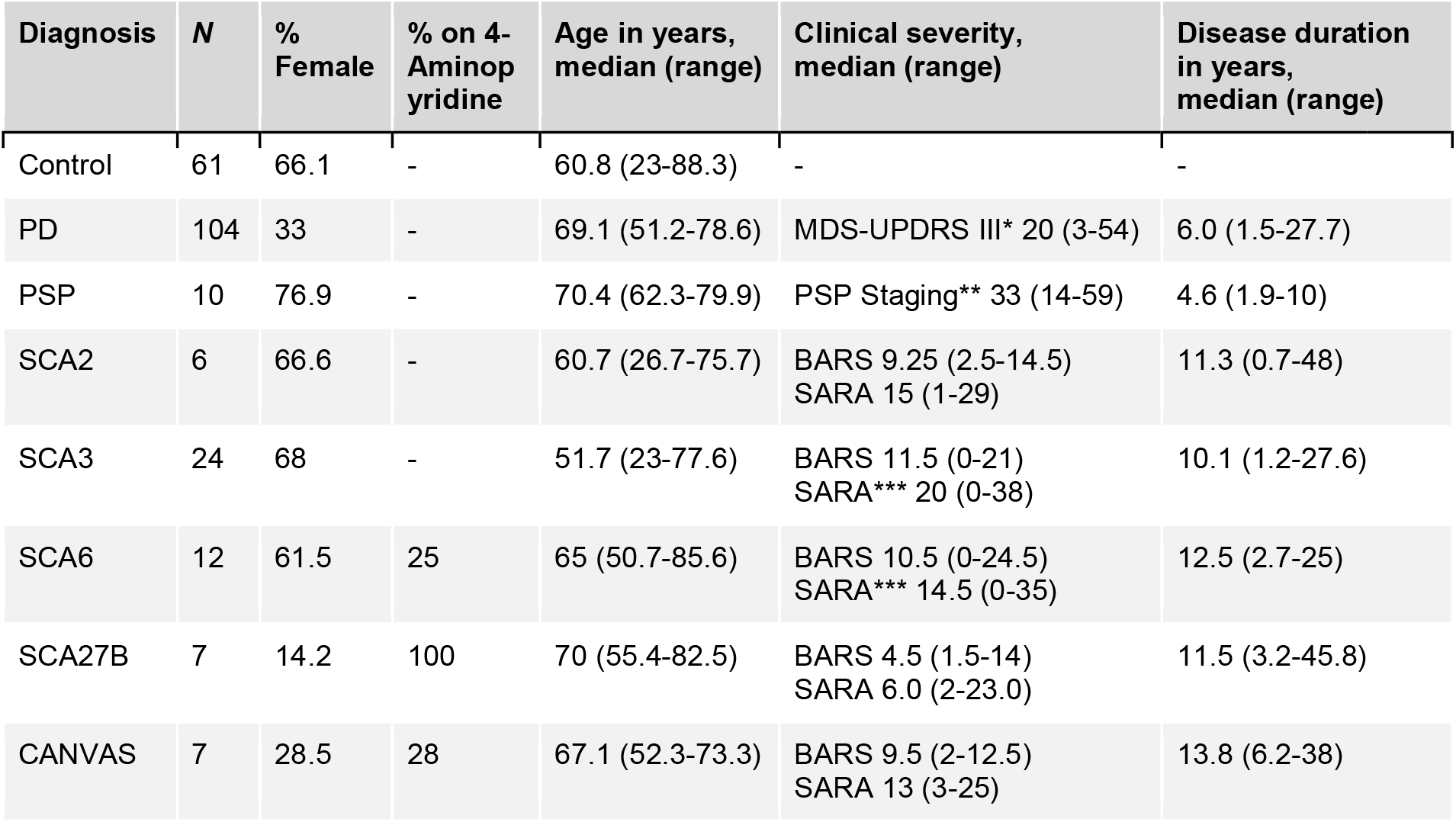
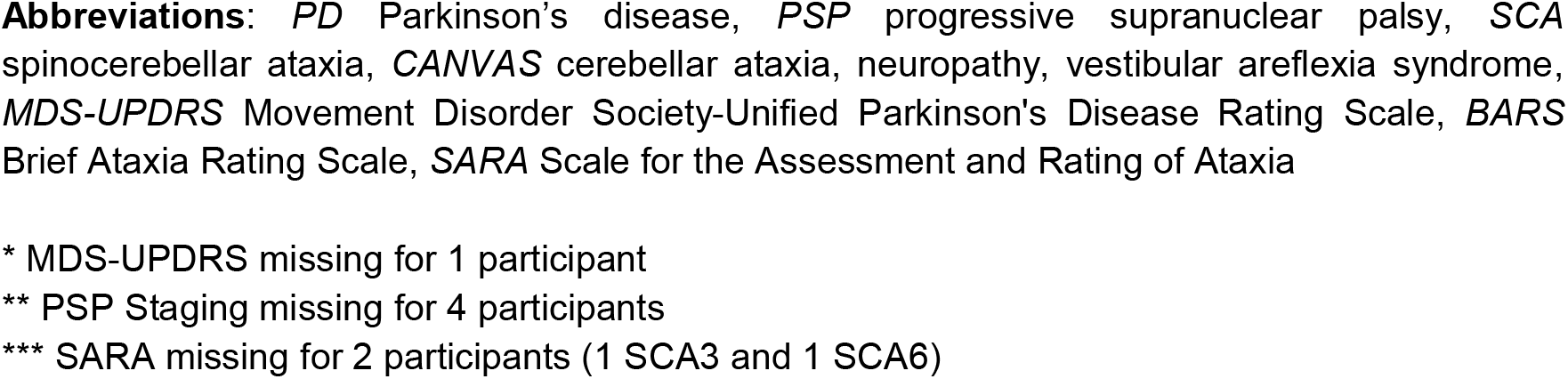
Participant Demographics – Disease Populations versus Controls Analysis.

| Diagnosis | N | % Female | % on 4-Aminopyridine | Age in years, median (range) | Clinical severity, median (range) | Disease duration in years, median (range) |
| --- | --- | --- | --- | --- | --- | --- |
| Control | 61 | 66.1 | - | 60.8 (23-88.3) | - | - |
| PD | 104 | 33 | - | 69.1 (51.2-78.6) | MDS-UPDRS III* 20 (3-54) | 6.0 (1.5-27.7) |
| PSP | 10 | 76.9 | - | 70.4 (62.3-79.9) | PSP Staging** 33 (14-59) | 4.6 (1.9-10) |
| SCA2 | 6 | 66.6 | - | 60.7 (26.7-75.7) | BARS 9.25 (2.5-14.5)<br>SARA 15 (1-29) | 11.3 (0.7-48) |
| SCA3 | 24 | 68 | - | 51.7 (23-77.6) | BARS 11.5 (0-21)<br>SARA*** 20 (0-38) | 10.1 (1.2-27.6) |
| SCA6 | 12 | 61.5 | 25 | 65 (50.7-85.6) | BARS 10.5 (0-24.5)<br>SARA*** 14.5 (0-35) | 12.5 (2.7-25) |
| SCA27B | 7 | 14.2 | 100 | 70 (55.4-82.5) | BARS 4.5 (1.5-14)<br>SARA 6.0 (2-23.0) | 11.5 (3.2-45.8) |
| CANVAS | 7 | 28.5 | 28 | 67.1 (52.3-73.3) | BARS 9.5 (2-12.5)<br>SARA 13 (3-25) | 13.8 (6.2-38) |

**Abbreviations:** *PD* Parkinson's disease, *PSP* progressive supranuclear palsy, *SCA* spinocerebellar ataxia, *CANVAS* cerebellar ataxia, neuropathy, vestibular areflexia syndrome, *MDS-UPDRS* Movement Disorder Society-Unified Parkinson's Disease Rating Scale, *BARS* Brief Ataxia Rating Scale, *SARA* Scale for the Assessment and Rating of Ataxia
\* MDS-UPDRS missing for 1 participant
\*\* PSP Staging missing for 4 participants
\*\*\* SARA missing for 2 participants (1 SCA3 and 1 SCA6)

The second analysis examined the relationship between VQoL and clinical oculomotor findings. This analysis was conducted for all individuals with ataxia and separately for a subset of individuals with SCA. The broader ataxia dataset included 167 sessions from 122 participants. The SCA subset included 85 sessions from 65 participants, including subtypes 1, 2, 3, 6, 8, 27B, and others. We excluded two presymptomatic individuals, who reported no symptoms related to their genetically confirmed ataxias, to reduce the cohort heterogeneity. Table 2 shows participant demographics in this analysis. Disease duration was calculated by subtracting the participants’ age at the onset of their first ataxia symptom from their age at the research visit.

**Table 2.** Ataxia Participant Demographics – Clinical Oculomotor Findings Analysis.

|  | All Ataxias (including SCAs) | SCAs Only |
| --- | --- | --- |
| *N* | 122 (total)<br>2 SCA-1, 5 SCA-2, 23 SCA-3,<br>11 SCA-6, 13 SCA-27B, 4 SCA-8,<br>7 other SCAs (types 7, 19/22, 28,<br>36)*<br>7 CANVAS, 6 FA, 2 FXTAS, 2<br>ARSACS<br>15 ANOS, 8 ARCA, 4 AIA, 4 DBN,<br>3 NPC, 1 VA, 1 EA, 1 SRA, 1 BCA,<br>1 GSS, 1 GHS, 1 NPCA | 65 (total)<br>2 SCA-1, 5 SCA-2, 23 SCA-3,<br>11 SCA-6, 13 SCA-27B, 4 SCA-8,<br>7 other SCAs<br>(types 7, 19/22, 28, 36)* |
| % Female | 55.5 | 60.6 |
| Age in years,<br>median (range) | 64.7<br>(21.3-85.6) | 65.4<br>(23-85.6) |
| Clinical severity,<br>median (range) | BARS 10.7 (0-24)<br>SARA** 16.5 (0-40) | BARS 10.5 (0-22.5)<br>SARA** 17 (0-35) |
| Disease duration in years,<br>median (range) | 11.8<br>(0.7-49.2) | 13<br>(0.7-49.2) |

**Abbreviations:** *SCA* spinocerebellar ataxia, *CANVAS* cerebellar ataxia, neuropathy, vestibular areflexia syndrome, *FA* Friedreich's ataxia, *FXTAS* Fragile X-associated tremor/ataxia syndrome, *ARSACS* autosomal recessive spastic ataxia of Charlevoix-Saguenay, *ANOS* ataxia, not otherwise specified, *ARCA* autosomal recessive cerebellar ataxia, *AIA* autoimmune-mediated ataxia, *DBN* downbeat nystagmus with ataxia, *NPC* Niemann-Pick disease type C, *VA* vestibular ataxia, *EA* episodic ataxia, *SRA* stroke-related ataxia, *BCA* Behcet's disease-associated cerebellar ataxia, *GSS* Gerstmann-Sträussler-Scheinker syndrome, *GHS* Gordon-Holmes syndrome, *NPCA* non-progressive cerebellar ataxia, *BARS* Brief Ataxia Rating Scale, *SARA* Scale for the Assessment and Rating of Ataxia
\* One participant with both SCA 6 and 8 is included in other SCAs
\*\* SARA missing for 3 participants (2 SCA3 and 1 SCA7)

### Patient-Reported Questionnaires

The Visual Activities Questionnaire (VAQ) is a 33-item instrument assessing vision-related quality of life in everyday tasks, including (Table 3 in Supplemental) It evaluates eight visual functions: color discrimination, glare disability, light/dark adaptation, visual acuity/spatial vision, depth perception, visual search, and visual processing speed (45). A 13-item subset most relevant to visual concerns in ataxias was selected, assessing visual acuity/spatial vision (4 items), depth perception (3 items), and visual processing speed (6 items). Responses were rated from 0 (“never”) to 4 (“always”), and composite scores were calculated for each domain and overall VQoL. Responses of “do not drive” were excluded for item 13 related to driving ability to ensure our analysis reflects participants’ current, active experience with driving. Participants were instructed to answer questions considering any visual aids they use, such as glasses or contacts. The original validation study reported mean VAQ total scores around 1.5 ± 0.5 in visually healthy adults (45). In clinical populations, a change of approximately 2.6 to 6.5 units in the VAQ score is considered clinically significant for patients with visual impairment (46).

**Table 3.** 13-item subset of the Visual Activities Questionnaire and pairwise Mann-Whitney U tests between all spinocerebellar ataxia individuals with and without nystagmus.

| Visual Function Question | p value | Cohen's d | 95% CI |
| --- | --- | --- | --- |
| Depth Perception |  |  |  |
| 9 - When pouring liquid, I have trouble judging the level of the liquid in a container, such as the level of coffee in a cup. | 0.026 * | 0.60 | (0.23, 1.10) |
| 21 - Sometimes when I reach for an object, I find that it is further away (or closer) than I thought. | 0.003 * | 0.76 | (0.35, 1.25) |
| 32 - I have problems judging how close or far things are from me. | 0.022 * | 0.52 | (0.04, 1.10) |
| Visual Acuity/Spatial Vision |  |  |  |
| 7 - I have problems reading small print (for example, phone book, newspapers). | 0.004 * | 0.74 | (0.35, 1.57) |
| 10 - I have trouble reading the menu in a dimly lit restaurant. | 0.013 * | 0.64 | (0.16, 1.38) |
| 15 - I have difficulty reading small print under poor lighting. | 0.003 * | 0.82 | (0.37, 1.63) |
| 27 - I have difficulty doing any type of work which requires me to see well up close. | 0.004 * | 0.73 | (0.33, 1.41) |
| Visual Processing Speed |  |  |  |
| 8 - I have trouble reading a sign or recognizing a picture when it's moving, such as an ad on a passing bus or truck. | 0.001 * | 0.84 | (0.49, 1.57) |
| 18 - I have problems carrying out activities that require a lot of visual concentration and attention. | 0.005 * | 0.66 | (0.20, 1.23) |
| 22 - I have difficulty noticing when the car in front of me is speeding up or slowing down. | 0.029 * | 0.47 | (0.01, 1.08) |
| 30 - When riding in a car, other cars on the road seem to be going too fast. | 0.238 | 0.20 | (-0.38, 0.90) |
| 33 - It takes a long time to get acquainted with new surroundings. | 0.014 * | 0.54 | (0.06, 1.01) |
| *13 - When I'm driving, other cars surprise me from the side, because I don't notice them until the last moment. | 0.023 * | 0.59 | (0.31, 2.05) |

\*Note: If a participant responds with "do not drive" on question 13, this item is excluded from the calculation.

Participants also completed the Patient-Reported Outcome Measure of Ataxia (PROM-Ataxia), a 70-item survey assessing physical function, daily living activities, and mental health (47). Three items in PROM-Ataxia are related to vision: 1) “I have double and/or blurred vision,” 2) “I have trouble with depth perception/judging distances,” and 3) “I am bothered by spinning sensations, dizziness, vertigo, or lightheadedness.” In both questionnaires, higher scores indicate lower quality of life.

### Clinical Rating Scales

Two clinical rating scales were performed by neurologists to evaluate four key clinical oculomotor signs: 1) abnormal eye movements at rest, 2) gaze-evoked nystagmus, 3) ocular pursuit abnormalities, and 4) dysmetria in saccades (overshoot or undershoot).

The BARS oculomotor subsection, scored in half-point increments from 0 to 2, provides an aggregate score based on the presence or absence of each sign, or if slowing is the prominent feature, an estimate of oculomotor slowing. The oculomotor component of the Modified International Cerebellar Ataxia Rating Scale (MICARS) additionally evaluates the severity of gaze-evoked nystagmus (rated as normal, transient, moderate, or severe) and ocular pursuit (rated as normal, slightly saccadic, or clearly saccadic) (43,48). For analysis, all MICARS items were coded as binary (0 for absent, 1 for present) to allow comparison with other oculomotor signs, which are not rated for severity.

To minimize temporal variability, VAQ responses more than 90 days from clinical assessments were excluded (10 out of the 167 sessions, or 5.9%).

### Statistical Analyses

Participant survey and clinical rating scale scores were analyzed using Python.

For the first analysis, we evaluated differences in VQoL between disease populations and controls. Mean and standard deviation for VAQ total score and each composite score were calculated for each disease group. Group differences were assessed using the nonparametric Mann-Whitney U test. The sample size (*n*=231) represents unique individuals. Controls were grouped by decade of life at the time of research participation. For each disease group versus control comparison, only age-matched controls were used. Age differences were statistically tested using the nonparametric Mann-Whitney U test. For the Parkinson’s disease analyses, we restricted controls to aged 50 to 80 to minimize confounding by age, as inclusion of younger controls resulted in significant age differences. The sample sizes for each disease population and their age-matched control group are reported in Figure 1. Effect size was reported using Cohen’s d with 95% confidence intervals.

**Figure 1.**
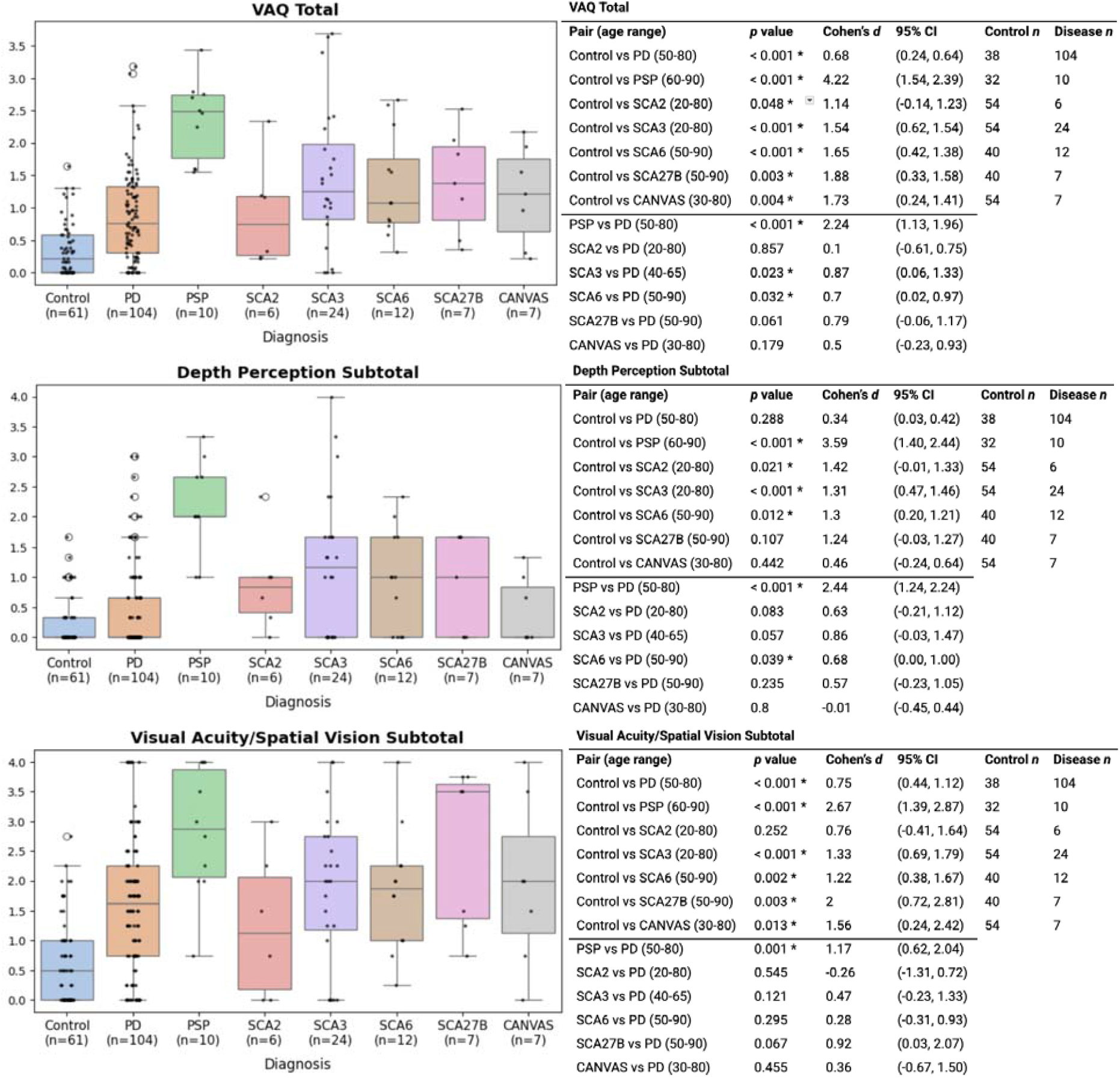
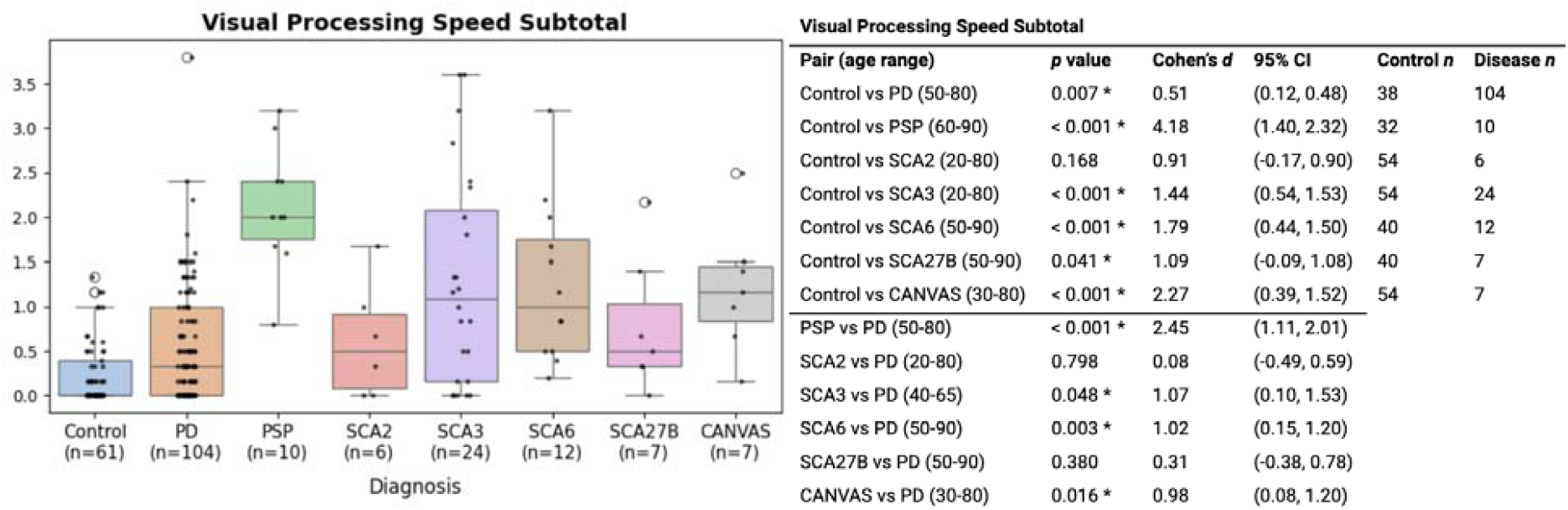
VAQ total (A), depth perception subtotal (B), visual acuity/spatial vision (C), and visual processing speed (D) as a function of diagnosis and respective pairwise Mann-Whitney *U* tests.

For the second analysis, we examined the relationship between patient-reported measures (VAQ and PROM-Ataxia) and severity of cerebellar oculomotor findings (BARS oculomotor subsection score) in ataxias using the Pearson correlation coefficient (*r*). The sample sizes (*n*=167 sessions from 122 participants) for all ataxias and (*n*=88 sessions from 65 individuals) for all SCAs reflect the total number of samples, including longitudinal data, from a set of unique individuals. The Mann-Whitney U test was used to compare VAQ scores for the presence or absence of each oculomotor abnormality (using binarized MICARS scoring). A regression model was run to predict VAQ total scores based on the four oculomotor signs assessed on MICARS.

### Data Sharing

Data can be requested by qualified researchers by visiting https://neurobooth.mgh.harvard.edu/.

## Results

### Disease Populations versus Controls

There were no differences in age between disease populations and control subsets, using the age-matching procedure described in Methods (*p*=0.334 for PD, *p*=0.669 for PSP, *p*=0.541 for SCA2, *p*=0.546 for SCA3, *p*=0.72 for SCA6, *p*=0.386 for SCA27B, and *p*=0.378 for CANVAS).

All disease populations reported significantly higher VAQ scores, indicating worse overall VQoL compared to controls (*p*<0.001, Cohen’s *d*=0.68 for PD; *p*<0.001, Cohen’s *d*=4.22 for PSP; *p*=0.048, Cohen’s *d*=1.14 for SCA2; *p*<0.001, Cohen’s *d*=1.54 for SCA3; *p*<0.001, Cohen’s *d*=1.65 for SCA6; *p*=0.003, Cohen’s *d*=1.88 for SCA27B; and and *p*=0.004, Cohen’s *d*=1.73 for CANVAS; Figure 1).

For PSP, SCA3, and SCA6, in addition to overall VAQ score, all subcategory scores were significantly worse compared to controls (Figure 1). In individuals with PD, visual acuity/spatial vision (*p*<0.001, Cohen’s *d*=0.75) and visual processing speed (*p*=0.007, Cohen’s *d*=0.51) were significantly different from controls but depth perception was not. Similar findings were observed in individuals with SCA27B (*p*=0.003, Cohen’s *d*=2 for visual acuity/spatial vision and *p*=0.041, Cohen’s *d*=1.09 for visual processing speed) and CANVAS (*p*=0.013, Cohen’s *d*=1.56 for visual acuity/spatial vision and *p*<0.001, Cohen’s *d*=2.27 for visual processing speed; Figure 1). In comparison, in SCA2 vs. controls, depth perception was significantly different (*p*=0.021, Cohen’s *d*=1.42) but visual acuity/spatial vision and visual processing speed were not.

### Parkinsonian Syndromes Comparison

Individuals with PSP reported significantly worse VQoL compared to individuals with PD across all visual function categories: depth perception (*p*<0.001, Cohen’s *d*=2.44), visual acuity/spatial vision (*p*=0.001, Cohen’s *d*=1.17), and visual processing speed (*p*<0.001, Cohen’s *d*=2.45; Figure 1).

### Ataxias Comparison

No significant differences were observed in overall VQoL or subcategories between the ataxia subtypes: SCA2, SCA3, SCA6, SCA27B, and CANVAS.

### Parkinson’s Disease vs. Ataxias

Given the large PD cohort, we used these individuals as a comparative population to assess the degree and relative pattern of visual impairment in ataxia subtypes. There were no differences in age between PD and ataxia subsets (*p*=0.331 for SCA2, *p*=0.627 for SCA6, *p*=0.538 for SCA27B, and *p*=0.389 for CANVAS) except SCA3 (*p*=0.015). For the PD vs SCA3 analysis, we restricted the comparison to individuals aged 40 to 65, given the challenge of aligning the age distributions between these populations. Importantly, SCA3 patients are generally younger and nevertheless exhibited worse VQoL, suggesting that age is unlikely to be a confounding factor in this comparison.

Individuals with SCA3 and SCA6 reported significantly worse overall VQoL compared to individuals with PD (*p*=0.023, Cohen’s *d*=0.87 and *p*=0.032, Cohen’s *d*=0.7, respectively; Figure 1). In individuals with SCA3, visual processing speed was significant (*p*=0.048, Cohen’s *d*=1.07), but depth perception and visual acuity/spatial vision were not. The same was observed in CANVAS (*p*=0.016, Cohen’s *d*=0.98). In individuals with SCA6, depth perception (*p*=0.039, Cohen’s *d*=0.68) and visual processing speed were significant (*p*=0.003, Cohen’s *d*=1.02), but visual acuity/spatial vision was not (Figure 1).

No significant differences were observed in overall VQoL or subcategories between individuals with SCA2 vs PD and SCA27B vs PD (Figure 1).

### Correlations with Clinical Oculomotor Findings in Ataxias

#### Univariate Analysis with VQoL PROMs

In the full ataxia cohort (*n*=167 sessions from 122 individuals), VAQ total showed a weak correlation with PROM-Ataxia world movement (*r*=0.35, *p*<0.001) and a moderate correlation with PROM-Ataxia double vision (*r*=0.55, *p*<0.001) and PROM-Ataxia total (*r*=0.59, *p*<0.001). A strong correlation was observed between VAQ total and PROM-Ataxia depth perception (*r*=0.71, *p*<0.001). BARS oculomotor total weakly correlated with VAQ total (*r*=0.24, *p*<0.01), VAQ depth perception (*r*=0.24, *p*<0.01), VAQ visual acuity/spatial vision (*r*=0.26, *p*<0.001), and VAQ visual processing speed (*r*=0.16, *p*<0.05; Figure 2).

**Figure 2.**
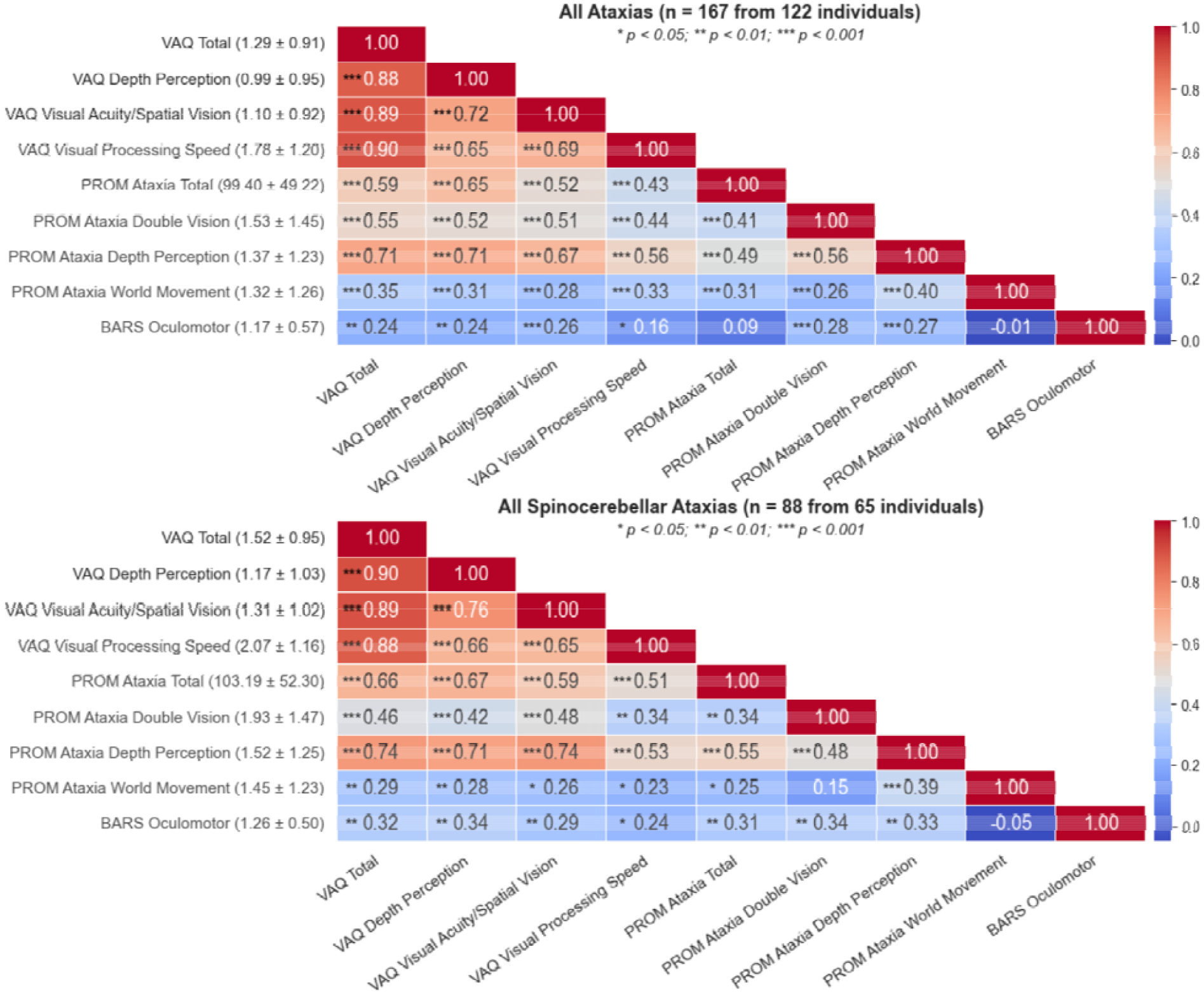
Pearson’s *r* correlations of patient-reported visual function with BARS oculomotor score in all ataxias (A) and all spinocerebellar ataxias (B). Statistical significance was indicated by an asterisk system (* *p* < 0.05; ** *p* < 0.01; *** *p* < 0.001).

In the SCA subset (*n*=88 sessions from 65 individuals), VAQ total and PROM-Ataxia items showed similar correlations (*r*=0.29, *p*<0.01 for PROM-Ataxia world movement, *r*=0.46, *p*<0.001 for PROM-Ataxia double vision, *r*=0.66, *p*<0.001 for PROM-Ataxia total, and *r*=0.74, *p*<0.001 for PROM-Ataxia depth perception). BARS oculomotor total showed slightly higher correlations with VAQ items, ranging from *r*=0.24 to *r*=0.34 (Figure 2).

#### Impact of Specific Oculomotor Signs on VQoL

To provide greater granularity in the assessment of specific oculomotor abnormalities on VQoL, we analyzed the binarized MICARS oculomotor variables. In the full ataxia cohort, the prevalence of each oculomotor sign was as follows: abnormal eye movement at rest (40/167), gaze-evoked nystagmus (101/167), saccadic pursuit (138/167), and dysmetria of saccade (98/167). Comparing VAQ total scores between individuals with and without each oculomotor sign, a significant association was found between the presence of nystagmus and saccadic pursuit and higher VAQ scores (*p*<0.001, Cohen’s *d*=0.65 and *p*=0.002, Cohen’s *d*=0.62 respectively; Figure 3). Abnormal eye movements at rest and dysmetria of saccade were not significant.

**Figure 3.**
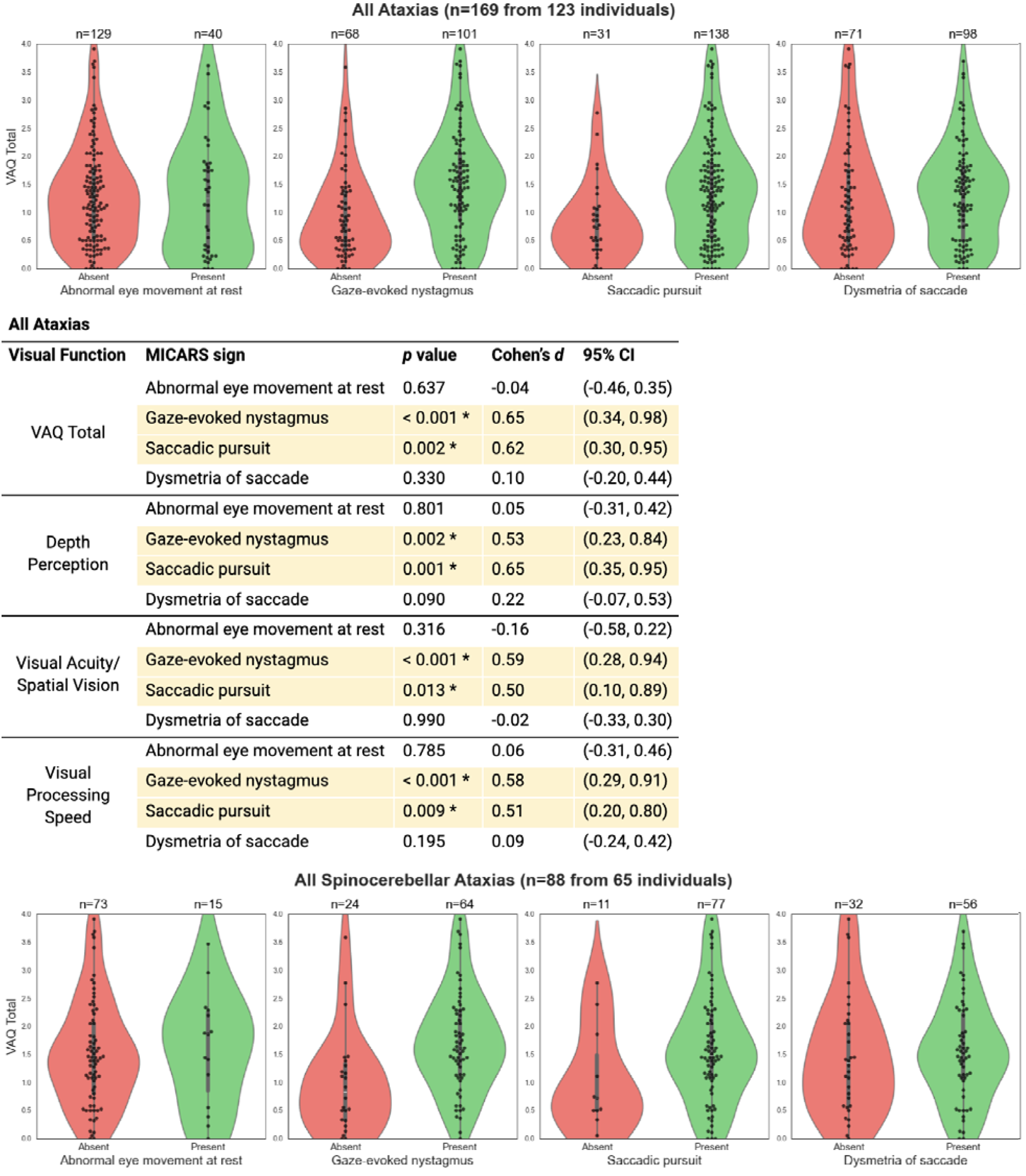
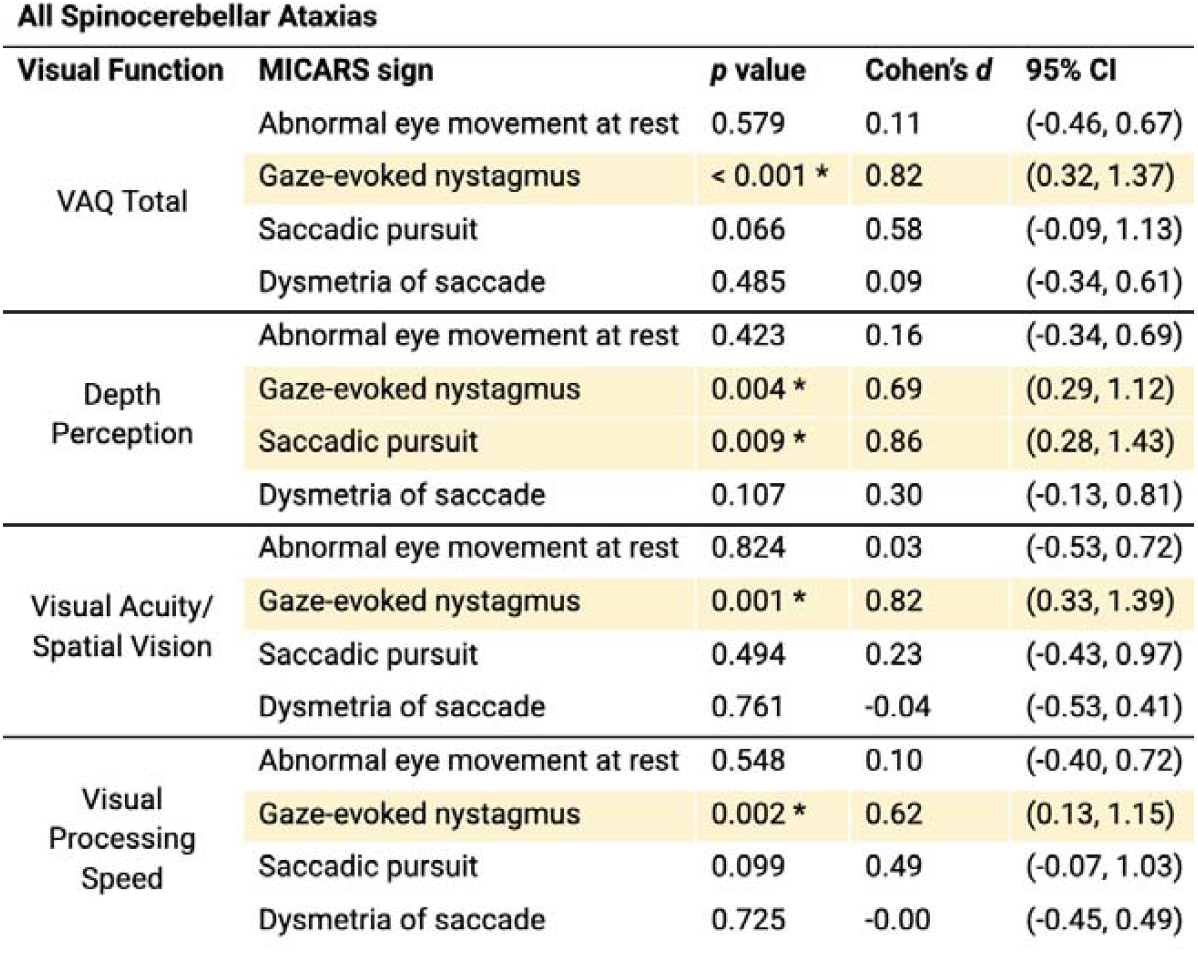
Distribution of MICARS oculomotor findings in all ataxias (A) and all spinocerebellar ataxias (B) and respective pairwise Mann-Whitney *U* tests.

In the SCA subgroup, the prevalence of each oculomotor sign was as follows: abnormal eye movement at rest (15/88), gaze-evoked nystagmus (64/88), saccadic pursuit (77/88), and dysmetria of saccade (56/88). Notably, in this subgroup, the presence of nystagmus was the only oculomotor sign associated with higher VAQ scores (*p*<0.001, Cohen’s d=0.82; Figure 3). Abnormal eye movements at rest, saccadic pursuit, and dysmetria of saccade were not significant.

#### Multivariate Regression with VAQ

A multivariate Ordinary Least Squares regression analysis was performed on both the all ataxias and all SCAs datasets to examine the combined influence of clinical oculomotor indicators on predicted VAQ total. The overall models were statistically significant but had limited predictive power (*p*<0.001, R^2^=0.159, adjusted R^2^=0.130 for all ataxias and *p*=0.029, R^2^=0.144, adjusted R^2^=0.094 for all SCAs).

Using the four oculomotor signs in binary form as independent variables for VAQ total, the only significant predictor of higher VAQ scores was the presence of nystagmus. This was observed in both the all ataxias (*p*=0.002) and all SCAs datasets (*p*=0.011) with a positive coefficient of 0.570 and 0.670, respectively. The other three oculomotor signs, abnormal eye movement at rest, saccadic pursuit, and dysmetria of saccade, were not significant predictors. This is consistent with the univariate analysis findings.

#### Pairwise Comparisons of Presence vs. Absence of Nystagmus

To further explore the relationship between nystagmus and VQoL, we examined specific VAQ items that differed between individuals with and without nystagmus in the SCA cohort. Individuals with nystagmus reported reduced VQoL in all 13 items, except for item 30 in the visual processing speed category: “When riding in a car, other cars on the road seem to be going too fast.” (*p*=0.238, Cohen’s *d*=0.20). The number of significant items for other signs is as follows: abnormal eye movements at rest (0/13), saccadic pursuit (4/13), and dysmetria of saccades (1/13).

The VAQ items that most significantly differentiated those with and without nystagmus were question 8 under visual processing speed: “I have trouble reading a sign or recognizing a picture when it’s moving, such as an ad on a passing bus or truck.” (*p*=0.001, Cohen’s *d*=0.84), question 15 under visual acuity/spatial vision: “I have difficulty reading small print under poor lighting.” (*p*=0.003, Cohen’s *d*=0.82), and question 21 under depth perception: “Sometimes when I reach for an object, I find that it is further away (or closer) than I thought.” (*p*=0.003, Cohen’s *d*=0.76; Table 3 in Supplemental).

## Discussion

In this study, we found that VQoL is strongly impacted across PD, PSP, and ataxia populations compared to age-matched controls, particularly in the visual processing speed domain. In evaluating relationships between VQoL and clinical oculomotor signs, gaze-evoked nystagmus emerged as a significant predictor of reduced VQoL in individuals with ataxia. Coarse/binary clinical oculomotor assessments only capture a small percentage of overall VQoL. These findings reinforce the importance of visual function in parkinsonian syndromes and ataxias, demonstrate the functional relevance of nystagmus, and support the use of additional tools in clinical care and clinical trials to evaluate visual function and visual quality of life.

Previous research has shown that individuals with Parkinson’s disease have impaired depth perception and visual processing speed (15,49–51) with medium to large effects sizes. Our results are consistent with these prior findings and extend knowledge about reduced visual acuity/spatial vision in PD. While visual acuity is typically believed to be preserved in the earliest stages of PD, with only mild reductions relative to age-matched controls (52), our findings suggest that broader aspects of visual function may be affected as the disease progresses.

Visual acuity as a measure can vary across studies depending on the parameters used. In our study, we asked participants to report VQoL with use of visual aids, reflecting their everyday experience. In PSP, we saw a significant reduction in all three vision domains with large effect sizes. This finding contrasts prior literature suggesting that oculomotor dysfunction in PSP does not usually cause lasting reductions in sharpness of vision (20). However, impairments in voluntary saccades and fixation control can disrupt visual exploration and reading, thereby affecting perceived visual function without altering static visual acuity (21). Additionally, secondary factors such as decreased blink rate can cause dryness, indirectly reducing vision clarity (53).

Prior work in ataxias has established the high prevalence of oculomotor abnormalities in spinocerebellar ataxias (34), including impaired depth perception in types 1, 3, and 6 (51,52) and impaired visual processing speed in types 1, 2, and 3 (53,54). This is consistent with prior studies comparing controls to SCA types 1, 3, and 6 (33) and SCA type 14 (54). Our results confirm these findings with a larger sample size with more SCA subtypes. While previous studies did not include effect sizes, we highlighted differences with consistently large Cohen’s *d*. We also present novel VQoL data from individuals with CANVAS and the recently described SCA27B (30). Compared to controls, both ataxia subsets showed reduced VQoL in visual acuity/spatial vision and visual processing speed. The largest differences were again observed in visual processing speed, with medium to large effect sizes, highlighting a magnitude of impairment not previously reported.

Importantly, we found a significant relationship between the presence of gaze-evoked nystagmus and overall VQoL, particularly in the domains of visual processing speed, and to a lesser extent, depth perception and visual acuity/spatial vision. Nystagmus can disrupt fixation stability, which is critical for reading and tracking moving objects. This instability likely contributes to the pronounced reductions in visual processing speed. Disrupted fixation may also result in blurred vision and oscillopsia, or illusory motion of the visual field, which can further diminish VQoL. The pronounced reduction in visual processing speed can also be associated with the presence of downbeat nystagmus, which is prevalent in SCA6, SCA27B, and CANVAS (55–57). Most studies of nystagmus and its impact of VQoL have focused on infant (58,59) and pediatric cases (60,61) with a gap in knowledge for adult neurodegenerative conditions.

It is also possible that nystagmus impacts VQoL more than other oculomotor signs because of its effect on stability during fixation, when most visual information processing is occurring (62– 64). In contrast, other symptoms, such as saccade or pursuit abnormalities, may less strongly relate to function because they do not occur throughout fixation and are less continuous or common during everyday behavior. This supports more granular quantification of the persistence and amplitude of nystagmus (and other oculomotor signs) to fully characterize their functional impact. Future work incorporating digital measures, especially during naturalistic behavior (65,66), may better explain individual differences in VQoL. At the bedside, the Scale for Ocular Motor Disorders in Ataxia can be used by clinicians (67) in combination with a nystagmus-specific quality of life instrument (68–70).

There were some limitations to the study. First, our findings relied, in part, on clinician-performed assessments, which provide a relatively coarse evaluation of each oculomotor sign. For instance, BARS uses a 0-2 scoring system, where a score of 1 can represent a variety of abnormal combinations, without accounting for the severity of each abnormality. Saccade speed, which is relevant in SCA2 and PSP, was not specifically assessed. The use of more objective and sensitive eye tracking technologies would offer more accurate detection and quantification. Our sample sizes were relatively small for SCA2 (*n*=6), SCA27B (*n*=7), CANVAS (*n*=7), and PSP (*n*=10). Given the rarity of these diseases, larger sample sizes are necessary to confirm study findings. With the large number of clinician-performed, patient-reported, and quantitative assessments in this natural history study we used a relevant subset of VAQ questions to reduce burden and support longitudinal retention. Additionally, while we included four questions related to visual acuity/spatial vision in the VAQ, we did not assess visual acuity using the gold-standard Early Treatment Diabetic Retinopathy Study chart (71). Vergence, which could impair depth perception was also not specifically assessed.

This study emphasizes the strong impact that parkinsonian syndromes and ataxias have on vision-related quality of life. In ataxias, clinical oculomotor assessments capture a relatively small proportion of visual quality life, thus patient-reported measures are essential and quantitative oculomotor assessments have potential to provide objective measures of function in the future.

## Data Availability

Data can be requested by qualified researchers by visiting https://neurobooth.mgh.harvard.edu/.

## Acknowledgment

The authors would like to thank the Neurobooth participants for their valuable time and perspectives.

## Authors’ Roles

Authors: Faye X. Yang, Rohin Manohar, Anna C. Luddy, Albert Y. Hung, MD, PhD, Anne-Marie A. Wills, MD, MPH, Christopher D. Stephen, MB, ChB, Jeremy D. Schmahmann, MD, Anoopum S. Gupta, MD, PhD

*List all authors along with a clarification of role(s): e*.*g. design, execution, analysis, writing, editing of final version of the manuscript*.

## Financial Disclosures of all authors (for the preceding 12 months)

### Financial Disclosure/Conflict of Interest

FXY, RM, ACL, AYH, CDS, and ASG have no relevant disclosures. AMW has received research funding from the NIH in conjunction with BioSensics. JDS discloses that he is the inventor of the PROM-Ataxia and Brief Ataxia Rating Scale studied in the manuscript, both of which are copyright-protected by The General Hospital Corporation.

### Funding Sources

This work was supported by NIH R01 NS117826, Massachusetts Life Sciences Center (MLSC), Biogen, Broad Institute, Dake Family Foundation, and Massachusetts General Hospital Department of Neurology.

## Financial Disclosures

### Financial Disclosure/Conflict of Interest

FXY, RM, ACL, AMW, AYH, CDS, and ASG have no relevant disclosures. JDS discloses that he is the inventor of the PROM-Ataxia and Brief Ataxia Rating Scale studied in the manuscript, both of which are copyright-protected by The General Hospital Corporation.

### Funding Sources

This work was supported by the NIH R01 NS117826.

## Supplementary Material

## Notes

Financial Disclosure/Conflict of Interest: FXY, RM, ACL, AMW, AYH, CDS, and ASG have no relevant disclosures. AMW has received research funding from the NIH in collaboration with BioSensics. JDS discloses that he is the inventor of the PROM-Ataxia and Brief Ataxia Rating Scale studied in the manuscript, both of which are copyright-protected by The General Hospital Corporation.

### Author Declarations

The study protocol was approved by the Massachusetts General Hospital Institutional Review Board (2021P000257).

